# Reframing “Paradoxical” Excitation: Disentangling EEG Complexity and Entropy Reveals Resting State Dynamics Associated with Propofol Susceptibility

**DOI:** 10.64898/2025.12.16.25342405

**Authors:** Derek Newman, Charlotte Maschke, George A. Mashour, Stefanie Blain-Moraes

## Abstract

**Background:** Propofol exposure can produce heterogenous neural responses, from the expected suppression to transient paradoxical excitation. EEG measures of signal complexity and entropy have emerged as reliable markers of consciousness, but different types of complexity and entropy measures are often conflated. We used Type I and II complexity measures on the Complexity–Entropy Causal Plane (CECP) to characterize divergent neural trajectories during propofol-induced loss of consciousness. We hypothesized that paradoxical excitation is reflected in both Type I and Type II complexity; that divergent trajectories on the CECP separate paradoxical excitation from suppression; and that baseline EEG complexity is associated with susceptibility to propofol.

**Methods:** We analyzed EEG data from two independent cohorts of healthy adults receiving propofol: the Chennu dataset (n = 20), which included resting-state baseline, mild, and moderate sedation, followed by a recovery period; and the RecCognition dataset (n = 8), which used escalating infusions from baseline to deep sedation. For each participant and sedation level, we extracted Lempel–Ziv Complexity (LZC; Type I) and Statistical Complexity (SC; Type II) and projected them onto the CECP. Pearson correlations related baseline SC to changes in SC during moderate sedation; behavioral responsiveness; effect-site propofol concentration; and time-to-loss-of-consciousness.

**Results:** At moderate sedation, participants who remained responsive showed paradoxical increases in LZC and decreases in SC, whereas unresponsive participants exhibited the opposite pattern. Baseline SC correlated negatively with both the change in SC (r = –0.88) and behavioural responsiveness, indicating that intrinsic brain dynamics influence individual susceptibility to sedation. CECP trajectories revealed a reproducible inflection point demarcating paradoxical excitation from suppression.

**Conclusions:** Mapping EEG trajectories on the CECP bridges anesthetic state transitions with underlying neural dynamics. Baseline neural complexity indexes individual sensitivity to propofol, determining whether brain dynamics transiently enter excitation or direct suppression.

## 1. Introduction

In the quest for neurophysiological markers of consciousness, temporal dynamics and information-theoretic measures are increasingly used to characterize brain signals. EEG signal complexity can track levels of consciousness^1^ across sleep^2,3^, general anesthesia^4–7^ and disorders of consciousness^8–10^. Using such measures, recent studies demonstrated a paradoxical increase in EEG complexity during anesthesia induction in some individuals^11–13^. Boncompte et al. (2021) reported increases in Lempel-Ziv complexity (LZC) during low dose propofol sedation^12^, recapitulating the often-observed biphasic effect of temporary behavioral and neurophysiological excitation during anesthesia induction^14–17^.

Importantly, not every person undergoing anesthetic induction demonstrates paradoxical excitation^12,17^, raising the question of the underlying reason for subject-specific heterogeneity in drug response. A potential explanation comes from the association of the brain’s baseline fluctuations to its response to perturbation^7,18–20^. Our team has demonstrated that pre-anesthetic brain dynamics relate to anesthetic-induced loss of complexity in patients in a disorder of consciosuness^21^. We have also shown that dynamic neural properties before a perturbation predict the evoked complexity following transcranial magnetic stimulation during exposure to propofol, xenon and ketamine anesthesia^7^.

To better describe the non-linearities during exposure to anesthesia, Sleigh and Hight (2021) advocated for complementing the widely used measure of LZC with measures of entropy and Type II complexity^17^. Briefly, complexity measures fall into two broad classes: Type I and Type II^22^. Type I measures, such as LZC, scale linearly with entropy, and are minimal in ordered systems and maximal at randomness. Type II measures follow an inverted U-shape relationship with entropy, which is maximal when systems balance order and randomness and structured patterns emerge (see ref.^23^ for a detailed discussion). Historically, complexity and entropy have often been conflated, yet they reflect different system properties: entropy quantifies randomness or unpredictability, whereas complexity reflects the degree of structure within that unpredictability. Type I and Type II complexities combine orthogonally in the Complexity-Entropy Causal Plane (CECP) framework^24,25^, producing a map of temporal dynamics that may reveal more nuanced trajectories of neural dynamics during exposure to anesthesia.

In this study, we investigate how EEG dynamics diverge between paradoxical excitation and suppression and identify dynamics which predispose individuals to specific trajectories in response to propofol. We first hypothesize that paradoxical effects are observable in both Type I and Type II complexity measures of EEG. We further hypothesize that paradoxical excitation is reflected in divergent trajectories of brain dynamics from wakefulness into anesthetic-induced unconsciousness on the CECP. Finally, we hypothesize that brain activity before drug exposure shapes susceptibility to anesthesia and individual drug responses, with more random brain activity associated with higher anesthetic susceptibility and lower paradoxical excitation.

## 2. Methods

### 2.1 EEG Data and Anesthesia Protocol

We retrospectively analyzed high-density EEG from two independent datasets. From Chennu et al. (2016) (hereafter, “Chennu dataset”), we analyzed the EEG of 20 healthy participants during four conditions (each 5 minutes): 1) baseline resting-state; 2) propofol mild sedation (0.6 μg/mL); 3) propofol moderate sedation (1.2 μg/mL); and 4) post-sedation resting-state^26,12^. Behavioral responsiveness was assessed using an auditory discrimination task throughout the protocol. Participants were classified as either “responsive” or “drowsy” based on a perceptual hit rate threshold of 60% during the moderate sedation condition (see Chennu et al, see Supplemental Methods). Drug concentration was defined as the estimated effect-site propofol concentration (in μg/mL), derived from blood samples collected each condition.

In the “RecCognition dataset”, 8 participants underwent a baseline EEG recording, followed by increasing propofol sedation of 100, 200, and 300 μg/kg/min (each 5 minutes), herein labeled as “baseline,” “light sedation,” “moderate sedation,” and “deep sedation” (see^27^ for the detailed protocol). Time-to-loss of consciousness (TTLOC) was defined as the time in seconds from the onset of propofol infusion to the last observed successful behavioral response (double hand-squeeze). This study is a subset of the Reconstructing Consciousness and Cognition (ReCCognition) study (NCT01911195), conducted at the University of Michigan Medical School and approved by the Institutional Review Board (HUM0071578).

### 2.2 Feature Extraction

Data was band-pass filtered between 0.5 and 45 Hz, resampled to 250 Hz, segmented into 10 second epochs and averaged referenced using MNE-Python^28^. To assess complexity across sedation states, we extracted LZC (Type I) and statistical complexity (SC) (Type II). LZC was computed using NeuroKit^29^ on the mean binarized amplitude envelope, derived from the analytic signal. SC was calculated using Ordpy^30^ on the continuous EEG signal. We aggregated EEG features per participant and per sedation state by computing the median across epochs and channels. For details on phase space reconstruction, see Supplemental Methods.

### 2.3 Statistical Analysis

We conducted all statistical analyses using Python and the SciPy^31^ and Statsmodels^32^ packages. For the Chennu dataset, Mann-Whitney U tests assessed differences in complexity features between baseline and moderate sedation within behavioral groups (responsive and drowsy), as well as between behavioural groups at each sedation condition.

To examine the relation between baseline complexity and response to propofol in the Chennu dataset, we performed Pearson correlations between baseline complexity and the change of complexity (e.g., Baseline – Moderate Sedation), perceptual hit rate performance, and propofol concentration during moderate sedation. These correlations were computed across the full sample and across responsive and drowsy groups. For the RecCognition data, a Pearson correlation was performed between baseline complexity and TTLOC. One participant was excluded for this analysis based on deviation from the trajectory of complexity across time (see Supplementary Materials).

### 2.4 Simulations

To interpret our results with respect to underlying dynamics, we generated simulated time series of regular oscillations, colored noise, and Fractional Brownian motion (fBm) signals, representing a range of deterministic and stochastic signals. We extracted SC (Type II) and PEn (Type I) complexity features for all simulated signals using Ordpy. We projected these values onto the CECP to create interpretable reference regions corresponding to distinct dynamical regimes—regular oscillations, colored noise, and fBm. We overlaid empirical data from the RecCognition dataset on this plane, visualizing trajectories in brain complexity dynamics from spontaneous wakefulness to pharmacologically induced unconsciousness. More information about the CECP is available in Supplemental Methods.

### 2.5 Identification of the inflection point

To identify the empirical inflection point in the Chennu dataset, we computed the change in Type II complexity (SC) from moderate to baseline sedation for each participant. We then conducted a Pearson correlation of these change scores against baseline SC values. The zero crossing of the best-fit regression line was taken as the complexity threshold, representing the inflection point that separates paradoxical excitation (decrease in SC) from suppression (increase in SC).

## 3. Results

### 3.1 Paradoxical effects of propofol are observable in both Type 1 and Type 2 complexity measures and vanish with higher concentrations of propofol

Replicating the key finding from Boncompte et al. (2021), who reported this effect using the Chennu dataset, LZC (i.e. Type I complexity) paradoxically increased in participants who remained behaviorally responsive during moderate sedation (U = 41.0, p = 0.027). In contrast, LZC decreased slightly in the drowsy group (i.e., participants who lost behavioral responsiveness) (U = 28.0, p = 0.710; Figure 1A). At moderate sedation, LZC was significantly higher in responsive compared to drowsy participants (U = 7.0, p = 0.001), with no group difference at baseline (U = 46.0, p = 1.0).

**Figure 1.**
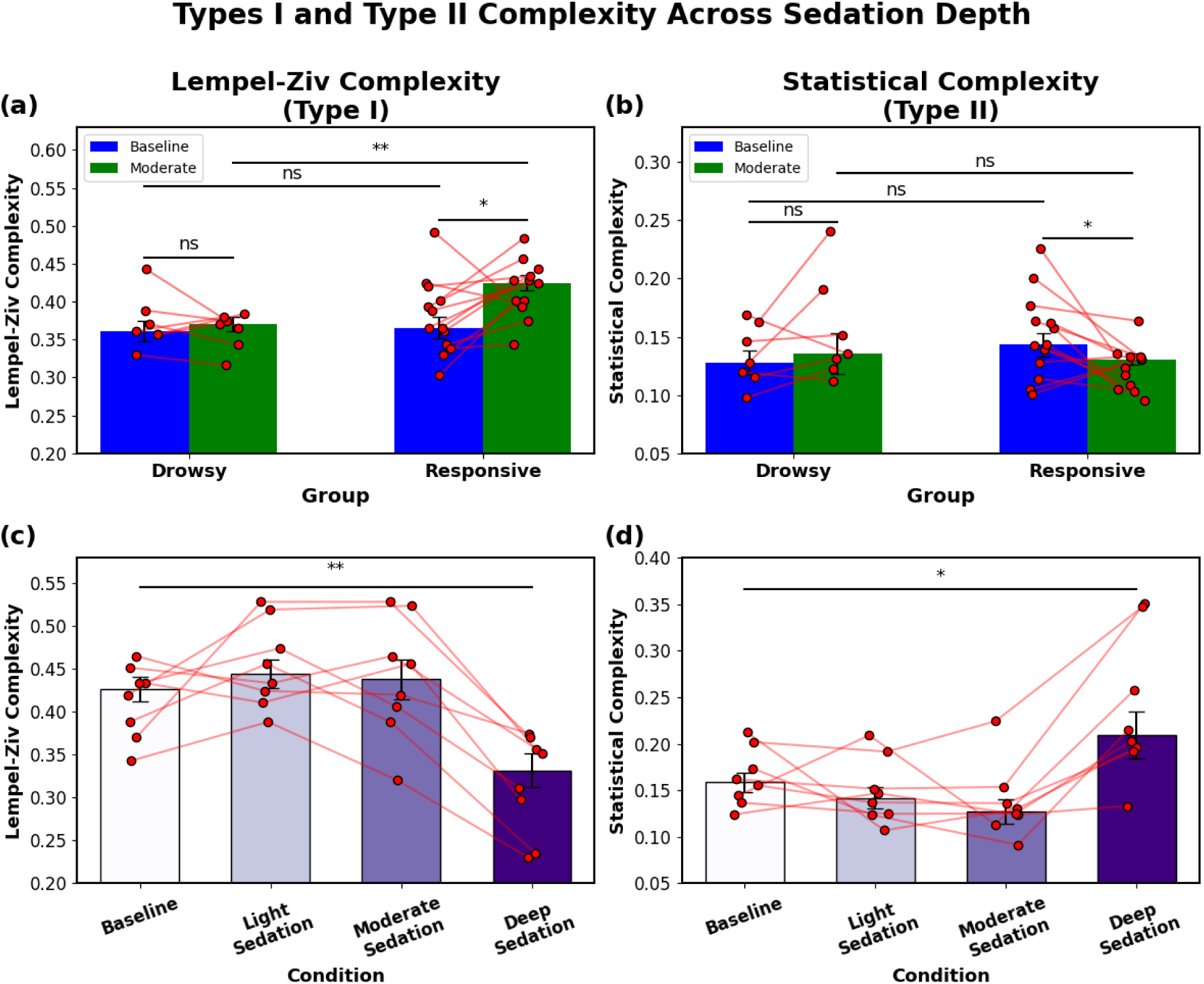
Type I and Type II complexity across sedation depth. (A) Lempel-Ziv Complexity (Type I) in the Chennu dataset shows a paradoxical increase from baseline to moderate sedation in participants who remained behaviorally responsive (green vs. blue), replicating Boncompte et al. (2021). No significant change was observed in drowsy participants. (B) Statistical Complexity (Type II) in the same dataset shows a significant decrease in responsive participants but no change in the drowsy group. (C) In the RecCognition dataset, Lempel-Ziv Complexity initially increases with mild and moderate sedation but significantly decreases at the deepest sedation level (Propofol 3), indicating a disappearance of paradoxical excitation. (D) Statistical Complexity follows an inverse trend: after a small reduction at lower doses, it significantly increases at Propofol 3, suggesting the emergence of reorganized neural dynamics during unconsciousness. Red dots and connecting lines represent individual participants. Statistical comparisons were assessed using Mann–Whitney U tests; significance thresholds are annotated as: *p < 0.05, **p < 0.01, ns = not significant.

SC significantly decreased from baseline to moderate sedation in the responsive group (U = 131.0, p = 0.018; Figure 1B), in the opposite direction of LZC. In contrast, the drowsy group showed a non-significant increase in SC (U = 18.0, p = 0.456). In high entropy neural signals, this opposite pattern—increase in Type I with decrease in Type II—is expected. Group differences in SC were not significant at either baseline or moderate sedation. It is important to note that LZC and SC are negatively related; our results are consistent with this relationship.

In the RecCognition dataset, LZC initially increased from baseline to light sedation, remained elevated at moderate sedation and then decreased sharply in deep sedation (U = 58.0, *p* = 0.005; Figure 1C). This decline coincided with the loss of behavioral responsiveness, which occurred for all participants during deep sedation. This non-linear profile mirrors the paradoxical excitation observed in the responsive group of the Chennu dataset. SC in the RecCognition dataset followed an inverse pattern—after a slight decrease at lower doses, SC increased from baseline to deep sedation (U = 13.0, *p* = 0.050; Figure 1D). All Mann–Whitney U test statistics are reported in Table 1.

**Table 1.** Summary of Mann–Whitney U tests comparing complexity measures across sedation conditions and behavioral groups. Each row reports group comparisons within or between sedation states for the first dataset and the second dataset. Complexity metrics include Lempel-Ziv Complexity (LZC, Type I) and Statistical Complexity (SC, Type II). Values shown include sample sizes (N), medians, standard deviations (STD), U statistics, p-values, and significance levels. Significance is denoted as follows: *p < 0.05, **p < 0.01, ns = not significant.

| DATASET | METRIC | GROUP 1 | GROUP 2 | N1 | MEDIAN1 | STD1 | N2 | MEDIAN2 | STD2 | U<br>STAT. | P-VAL. | SIG. |
| --- | --- | --- | --- | --- | --- | --- | --- | --- | --- | --- | --- | --- |
| 1 | LZC | Drowsy<br>Baseline | Drowsy<br>Moderate | 7 | 0.361 | 0.036 | 7 | 0.37 | 0.024 | 24.5 | 1.000 | ns |
| 1 | LZC | Responsive<br>Baseline | Responsive<br>Moderate | 13 | 0.366 | 0.049 | 13 | 0.424 | 0.036 | 38.5 | 0.019 | * |
| 1 | LZC | Drowsy<br>Baseline | Responsive<br>Baseline | 7 | 0.361 | 0.036 | 13 | 0.365 | 0.049 | 40.5 | 0.721 | ns |
| 1 | LZC | Drowsy<br>Moderate | Responsive<br>Moderate | 7 | 0.37 | 0.024 | 13 | 0.424 | 0.036 | 8.0 | 0.003 | ** |
| 1 | SC | Drowsy<br>Baseline | Drowsy<br>Moderate | 7 | 0.128 | 0.026 | 7 | 0.136 | 0.045 | 18.0 | 0.456 | ns |
| 1 | SC | Responsive<br>Baseline | Responsive<br>Moderate | 13 | 0.143 | 0.036 | 13 | 0.130 | 0.018 | 124.0 | 0.046 | * |
| 1 | SC | Drowsy<br>Baseline | Responsive<br>Baseline | 7 | 0.128 | 0.045 | 13 | 0.143 | 0.036 | 35.0 | 0.438 | ns |
| 1 | SC | Drowsy<br>Moderate | Responsive<br>Moderate | 7 | 0.136 | 0.045 | 13 | 0.131 | 0.018 | 66.0 | 0.115 | ns |
| 2 | LZC | Baseline | Propofol 3 | 8 | 0.427 | 0.042 | 8 | 0.3332 | 0.058 | 58.5 | 0.006 | ** |
| 2 | SC | Baseline | Propofol 3 | 8 | 0.159 | 0.031 | 8 | 0.21 | 0.077 | 12.0 | 0.038 | * |

In summary, we replicated the paradoxical increase in LZC during moderate sedation reported by Boncompte et al., (2021) and extended this by showing that Type II complexity (SC) exhibits a complementary pattern—decreasing at moderate sedation and increasing at deep sedation. These results demonstrate that both Type I and II complexity measures capture paradoxical excitation that is only expressed at moderate sedation and vanishes during deep sedation.

### 3.2 Baseline EEG complexity is associated with individual neural and behavioral responses to sedation

Having established that both Type I and Type II complexity measures track group-level changes with sedation, we next focus on whether baseline Type II brain complexity is associated with individual neural and behavioral responses to propofol.

#### Neural response

In the Chennu dataset, the change in SC from baseline to moderate sedation was significantly negatively correlated with baseline SC (r = –0.70, p < 0.001; Figure 2A), indicating that higher baseline SC yields stronger loss of SC during drug exposure. This effect was strongest in the responsive subgroup (r = –0.89, p < 0.001) and absent in the drowsy group (r = – 0.19, p = 0.68). This relationship was replicated in the RecCognition dataset, where baseline SC was associated with the reduction in SC at moderate sedation (r = –0.81, p = 0.028; Figure 2B), demonstrating that the neurophysiological response to propofol is directly associated with baseline SC.

**Figure 2.**
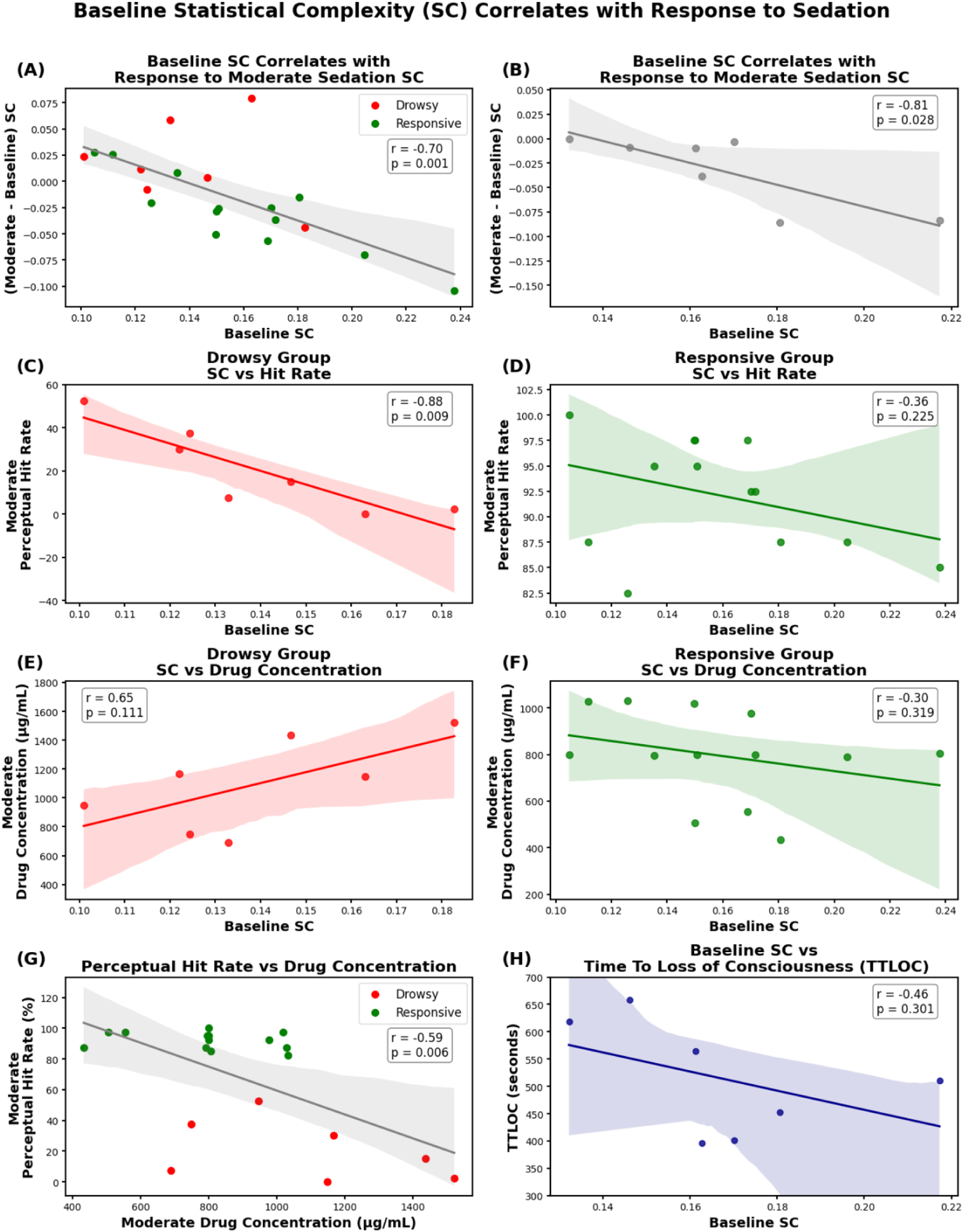
Baseline complexity is associated with neural and behavioral sensitivity to sedation. (A) In the Chennu dataset, higher baseline statistical complexity (Type II) was associated with larger changes in complexity during moderate sedation. This relationship was strongest in participants who remained behaviorally responsive (green) and absent in those who became drowsy (red). (B) This pattern replicated in a RecCognition dataset using a moderate propofol dose (Propofol 2), confirming that baseline complexity is associated with the magnitude of neural response to sedation. (C–D) Baseline complexity also reflects behavioral responsiveness. In the drowsy group (C), greater complexity was associated with lower perceptual hit rates under sedation (r = –0.88, p = 0.009), whereas the responsive group (D) showed a similar trend that did not reach significance. (E–F) Baseline complexity was not significantly correlated with drug concentration in either group, though the trends were in opposite directions—suggesting that brain state, not pharmacokinetics, drives sedation response. (G) Drug concentration significantly reflects hit rate across participants, highlighting its pharmacodynamic effect on responsiveness. (H) In the RecCognition dataset, higher baseline complexity was associated with faster time to loss of consciousness (TTLOC). Shaded regions denote 95% confidence intervals of linear fits; statistics reflect Pearson correlation coefficients.

#### Behavioral response

Baseline SC was also associated with behavioral responsiveness. In the drowsy group, baseline SC correlated with perceptual hit rates during the task under moderate sedation (r = – 0.88, p = 0.009; Figure 2C), explaining 77% of the variance. Here, greater baseline SC was associated with reduced responsiveness during drug exposure, indicating a suppressive sedation effect. No significant relationship was observed in the responsive group (r = –0.36, *p* = 0.23; Figure 2D). Thus, in individuals whose command-following was impaired by propofol, higher baseline SC indicates a greater behavioral suppression under sedation, whereas in responsive participants, higher baseline SC corresponds to paradoxical increases in neural activity (Figure 2A).

#### Relationship to drug concentration

Crucially, these effects were not explained by differences in drug concentration. While drug concentration and hit rate during moderate sedation was significantly correlated across participants (r = –0.59, *p* = 0.006; Figure 2G), baseline SC was only weakly correlated with drug concentration (Drowsy: r = 0.65, *p* = 0.11; Responsive: r = –0.30, *p* = 0.32; Figures 2E–F), and in opposite directions for each group. This dissociation suggests that SC-responsiveness relationships reflect a functional property of the intrinsic neural state rather than pharmacokinetics.

#### Time to loss of consciousness

In the RecCognition dataset, greater baseline SC was also associated with shorter time to loss of consciousness (r = –0.46, *p* = 0.30; Figure 2H). Although not statistically significant, this trend suggests that baseline SC may reflect susceptibility to anesthesia, with high-SC brain states more susceptible to rapid transitions under pharmacological perturbation. To demonstrate the robustness of these findings, we reproduced Figure 2 using LZC (Type I), where comparable but weaker associations were observed (see Supplementary Results).

Taken together, we demonstrate that baseline EEG complexity—particularly Type II complexity—is associated with both neural and behavioral sensitivity to sedation. These effects were not explained by drug concentration, underscoring the role of intrinsic brain states as an indicator of individual response to sedation. All correlation statistics are presented in Table 2. However, while these findings reveal a strong relation between baseline complexity and the perturbational response, they do not distinguish whether these changes result in suppression or paradoxical excitation. We therefore map the brain states in the CECP.

**Table 2.** Correlation statistics between baseline complexity, behavioral responsiveness, drug concentration, and loss of consciousness. Summary of group size (n), Pearson correlations (r), p-values, and variance explained (r²) for the relationships between baseline EEG complexity and neural or behavioral responses to sedation across both datasets. Group-wise correlations for Panels A and G are also presented separately (bottom rows). Significance is denoted as follows: ns = not significant, *p < 0.05, **p < 0.01, ***p < 0.001.

| *Panel* | *Variables* | *n* | *r* | *p* | *r*<sup>2</sup> | *Significant* |
| --- | --- | --- | --- | --- | --- | --- |
| *A* | Baseline Complexity vs ΔComplexity (Moderate) | 20 | -0.6958 | 0.0007 | 0.4841 | *** |
| *B* | Baseline Complexity vs ΔComplexity (Propofol2) | 7 | -0.8078 | 0.0280 | 0.6526 | * |
| *C* | Baseline Complexity vs Hit Rate (Drowsy) | 7 | -0.8801 | 0.0090 | 0.7745 | ** |
| *D* | Baseline Complexity vs Hit Rate (Responsive) | 13 | -0.3616 | 0.2247 | 0.1308 | n.s. |
| *E* | Baseline Complexity vs Drug Conc. (Drowsy) | 7 | 0.6539 | 0.1111 | 0.4275 | n.s. |
| *F* | Baseline Complexity vs Drug Conc. (Responsive) | 13 | -0.3005 | 0.3185 | 0.0903 | n.s. |
| *G* | Drug Conc. vs Hit Rate (All) | 20 | -0.5891 | 0.0063 | 0.3470 | ** |
| *H* | Baseline Complexity vs TTLOC | 7 | -0.4581 | 0.3013 | 0.2098 | n.s. |
| *A_Drowsy* | Baseline Complexity vs ΔComplexity (Moderate) | 7 | -0.1941 | 0.6767 | 0.0377 | n.s. |
| *A_Responsive* | Baseline Complexity vs ΔComplexity (Moderate) | 13 | -0.8856 | 0.0001 | 0.7844 | *** |
| *G_Drowsy* | Drug Conc. vs Hit Rate | 7 | -0.4037 | 0.3691 | 0.1630 | n.s. |
| *G_Responsive* | Drug Conc. vs Hit Rate | 13 | -0.2478 | 0.4143 | 0.0614 | n.s. |

### 3.3 Mapping the response to anesthesia on a complexity-entropy causal plane reveals a critical inflection point

To interpret results on the CECP, we first projected simulated signals representing distinct dynamical regimes—regular periodic signals with different frequency components, colored noise with varying spectral slopes, and fractional Brownian motion with different Hurst exponents—alongside experimental neural signals. These simulations each reflect fundamental properties of deterministic and stochastic processes (see Supplementary Results for details).

Superimposing empirical EEG data from the RecCognition dataset on the CECP reveals a structured degradation pathway towards propofol-induced unconsciousness. Group-averaged EEG trajectories (Figure 3) trace a distinct, non-linear path: from intermediate complexity and entropy baseline (blue dot), through mild and moderate sedation (yellow and green), where complexity decreases and entropy increases, to deep sedation (red) which shows a reversal of higher complexity and lower entropy. This group pattern closely matches individual participant trajectories (Figure 4C). Notably, moderate sedation occupies a region of decreased complexity and increased entropy, representing a transient, paradoxical state of heightened disorder.

**Figure 3.**
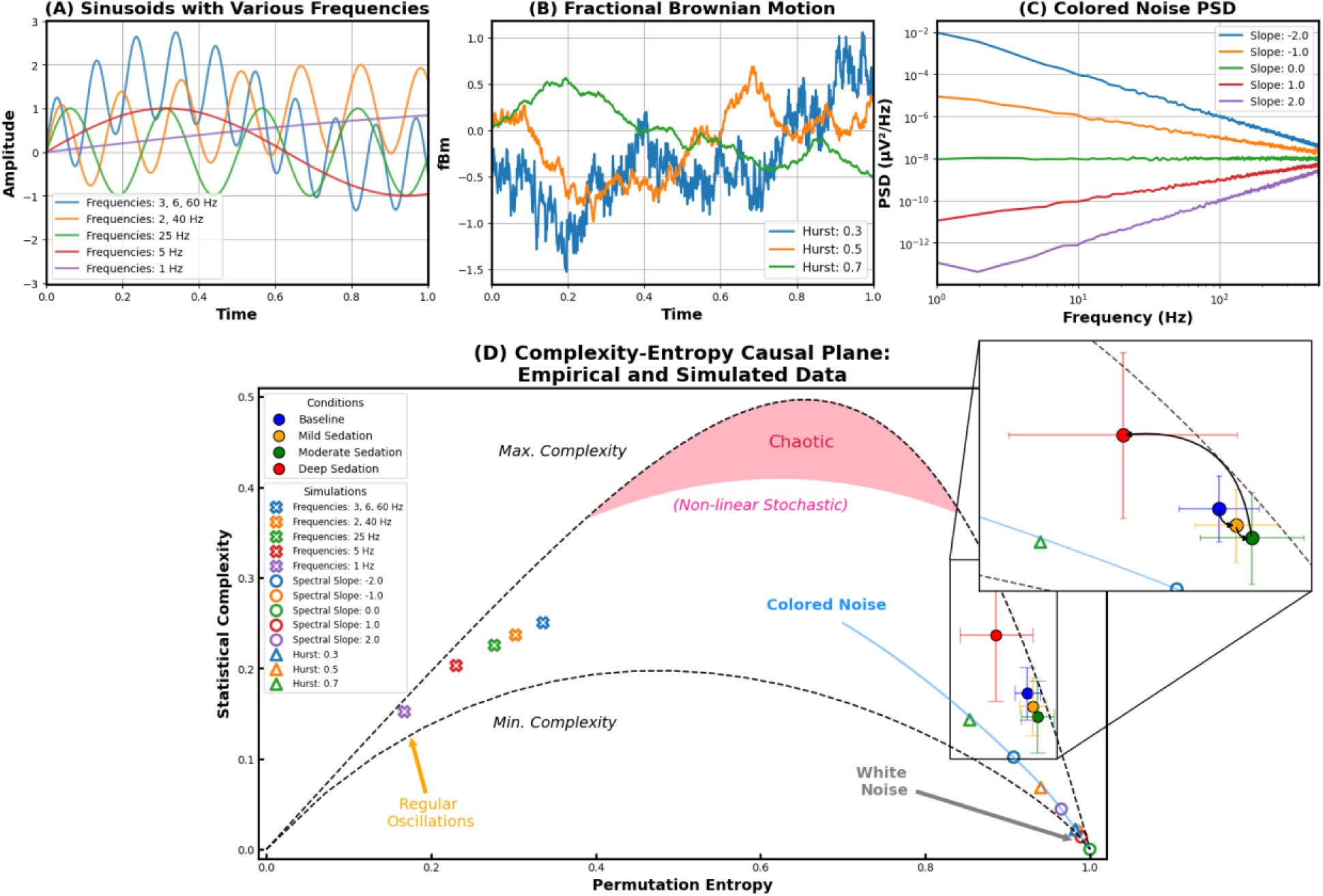
Complexity–Entropy Causal Plane reveals non-linear brain dynamics under sedation. (A)–(C) Simulated signals benchmark key dynamical regimes: (A) deterministic sinusoids of varying frequencies (denoted by crosses); (B) fractional Brownian motion (Hurst exponents H = 0.3, 0.5, and 0.7) (denoted by circles); and (C) colored noise spectra with power-law slopes (–2 to +2) (denoted by triangles). (D) The Complexity–Entropy Causal Plane (CECP) maps empirical data from the second dataset (Baseline (blue), Propofol 1–3 (yellow, green, and red)) alongside simulations. Black arrows depict state trajectories of group-average brain states under escalating sedation, starting at baseline and ending at the deepest sedation (Propofol 3). Background regions demarcate theoretical limits: the upper and lower envelope defines maximal and minimal statistical complexity, chaotic region in (red) and colored stochastic (blue) regimes. Adapted from Zanin & Olivares (2021).

**Figure 4.**
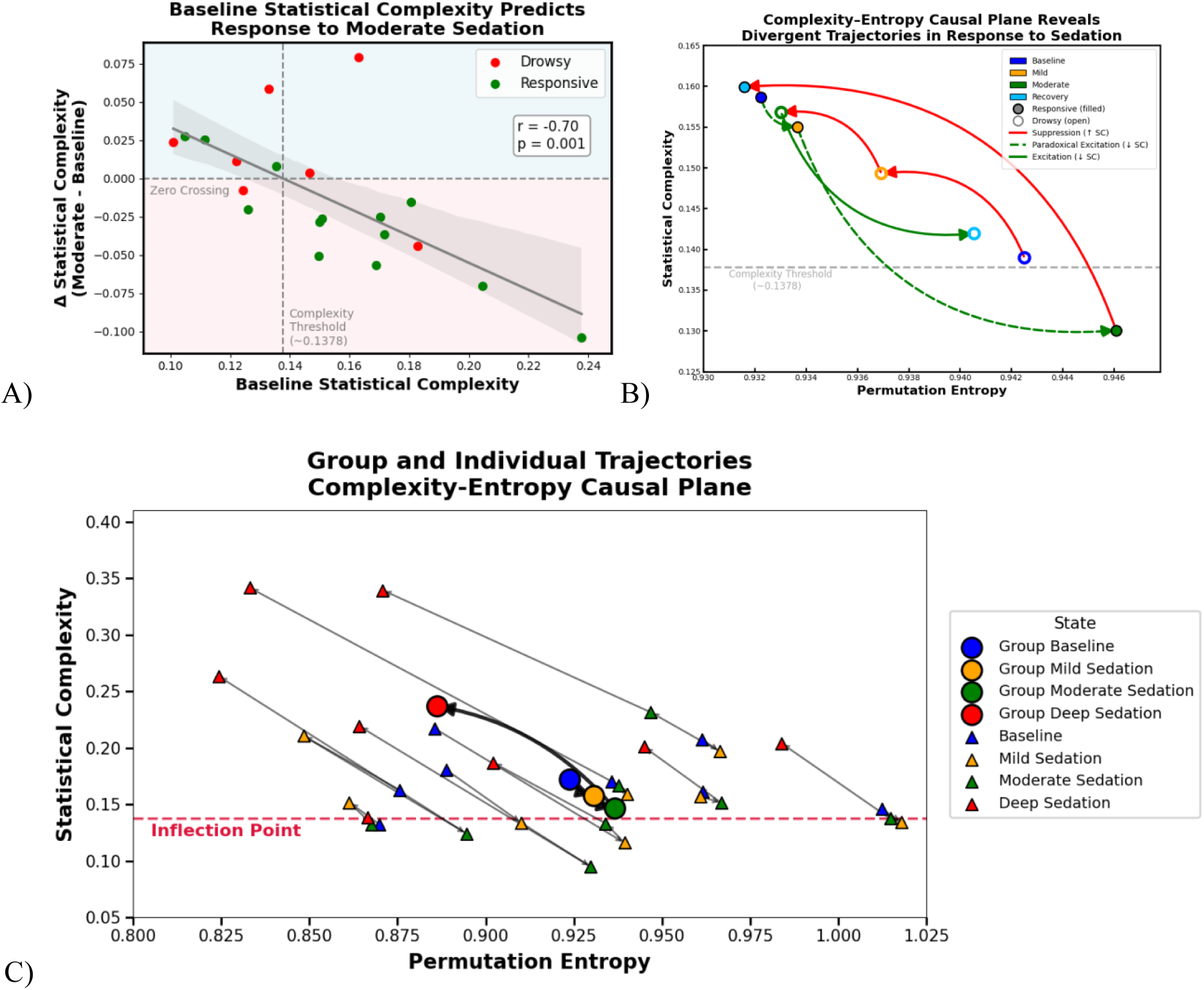
Statistical complexity defines an inflection point across both datasets that explains divergent trajectories to unconsciousness. (A) In the Chennu dataset, the inflection point (“complexity threshold”) was identified from the point where the regression line crossed zero change in complexity from baseline to moderate sedation. This value (∼0.1378) represents the moderate sedation complexity at which baseline brain dynamics are most similar across participants. Individuals above this threshold (light blue region) tended toward suppression (increased complexity), whereas those below it (light pink region) tended toward paradoxical excitation (decreased complexity). (B) In a zoomed-in Complexity–Entropy Causal Plane (CECP), group trajectories diverge according to sedation response. Responsive participants (solid markers) show decreased complexity (paradoxical excitation) at moderate sedation, whereas drowsy participants (open markers) show monotonic increases in complexity (suppression) during sedation. Red arrows indicate suppression, dashed green arrows indicate paradoxical excitation, and solid green arrows indicate excitation. (C) A zoomed-in CECP panel plots individual participant (triangles) trajectories from the RecCognition dataset across each condition (Baseline (blue), Mild (yellow), Moderate (green), and Deep Sedation (red)), with jitter applied for visibility. Large circles show group means for each state. Grey and black arrows indicate the progression from baseline to deep sedation for both individuals (thin) and group means (thick). The dashed red line marks the empirically observed inflection point derived from the Chennu Dataset (detailed in methods). Individual trajectories closely follow the group average, highlighting consistent shifts in brain dynamics during sedation.

This progression to unconsciousness results in a distinct inflection point at moderate sedation, where the trajectory shifts direction as complexity first decreases, then increases sharply during deep sedation (i.e., transitions from paradoxical excitation into anticipated suppression). The trajectory of every participant passes through this inflection point. While previous results showed that baseline complexity was associated with the response to sedation, the CECP reveals the direction of this change and how propofol sedation alters the balance between order and disorder in neural activity.

### 3.4 The inflection point explains divergent trajectories under sedation

While baseline EEG complexity is associated with the magnitude of response to sedation (Figure 2A), it does not directly reveal the direction of that change—whether an individual’s brain became paradoxically more excited or suppressed. We identified an empirical inflection point from the Chennu dataset. In Figure 4A (see also Figure 2A), the magnitude of response during moderate sedation is shown; individuals with lower baseline SC showed increases in SC, while those with higher baseline SC exhibited decreases in SC. The point which separates responsive and drowsy participants (i.e., the zero crossing of this fit) marks a complexity threshold (∼0.1378)—a data-driven estimate of the boundary between paradoxical excitation and suppression. This inflection point partitions individuals: those below the threshold (light pink), who exhibit paradoxical excitation, and those above (light blue), who exhibit suppression.

Projecting both the drowsy and responsive subgroups’ trajectories across sedation levels onto the CECP (Figure 4B) reveals a shared transitional zone—an intermediate region of high entropy and low complexity that all participants pass through. Beyond this zone, trajectories diverge. The responsive group starts with higher baseline complexity (further from the inflection point) and moves into a regime of paradoxical excitation (decreased complexity, increased entropy). In contrast, the drowsy group starts closer to the threshold, transitions into a suppressed regime (increased complexity, decreased entropy) marking a loss of responsiveness and consciousness. Baseline complexity thus acts as a neural dynamic marker of susceptibility to anesthesia: the proximity of baseline complexity to this inflection point determines an individual’s response to sedation—those at greater distance will maintain responsiveness, and those at smaller distances are easily nudged to unconsciousness.

This complexity threshold derived from the Chennu dataset closely matches the inflection point observed from the CECP plane with the RecCognition dataset (Figure 4C). This consistency strengthens the interpretation that the inflection point represents a stable boundary in brain state-space separating paradoxical excitation from suppression.

Taken together, these results show that the CECP not only maps neural state transitions but also captures the divergent dynamics underlying sedation response. This identification of the inflection point with the CECP provides a principled framework for interpreting divergent outcomes (paradoxical excitation vs. suppression) based on relative position to the inflection point before exposure to anesthesia.

## 4 Discussion

Across two independent datasets, we show that Type I and Type II EEG complexity capture distinct yet complementary aspects of the brain’s response to anesthesia. EEG complexity before exposure to propofol is associated with both neural and behavioral drug susceptibility. This effect was independent of drug concentration, indicating that response heterogeneity result from intrinsic neural dynamics before drug exposure. Baseline complexity thus serves as a neural marker of anesthesia susceptibility, capturing the system’s initial condition approaching bifurcation. Projecting trajectories of pharmacologically induced unconsciousness onto the CECP reveals a reproducible inflection point that underpins distinct trajectories separating paradoxical excitation from suppression. Paradoxical excitation thus reflects the interaction between baseline complexity and the inflection point where endogenous temporal structure collapses before exogenous inhibitory drug effects impose deterministic constraints.

Boncompte et al. (2021) introduced a susceptibility-to-propofol index that normalized behavioral loss by drug concentration and showed it correlated with LZC changes, defining susceptibility as dose sensitivity. In contrast, our findings highlight baseline brain complexity itself as the substrate of susceptibility—linking intrinsic neural dynamics to responsiveness. This perspective echoes the Fluctuation–Dissipation Theorem, which posits that a system’s response to perturbation is proportional to its baseline fluctuations^18–20^.

The CECP has previously been applied to experimental neural data, from EEG sleep stages^33^, discrimination of imagined versus executed motor tasks^34^, and MEG recordings in mild cognitive impairment and Alzheimer’s disease^35^. In our study, projecting EEG trajectories onto the CECP revealed a reproducible, non-linear pathway from baseline to unconsciousness, with all individuals passing through a shared noisy intermediate region during sedation. This region—low SC and high entropy—marks a transient state of paradoxical excitation. Empirical trajectories overlapped with colored noise simulations, but showed higher SC across states, indicating preserved temporal dependencies. SC at first decreases, reflecting loss of endogenous temporal structure before rebounding as drug effects imposed deterministic constraints. This result supports the entropic brain hypothesis that pharmacological perturbations can push the brain into higher-entropic states^36^.

Our results position baseline complexity as a meaningful indicator of anesthesia sensitivity. The degree of susceptibility depends on how much endogenous temporal structure the system holds at baseline. Brains which are already closer to the unstable regime are more easily tipped into suppression, whereas less random and more ordered brains sustain paradoxical excitation longer before collapse. This interpretation helps explain why individual state factors such as fatigue or stress—both of which alter baseline temporal structure—can shape anesthesia responsiveness.

The results of this study need to be interpreted within the limitations of low sample size and the exclusive use of propofol anesthesia. Future work should test larger cohorts with standardized anesthetic protocols, real-time CECP tracking, and application across anesthetic agents and clinical populations. The ability to map individual trajectories toward unconsciousness has clear clinical relevance. Baseline SC and CECP positioning could guide personalized anesthetic titration and perioperative monitoring, reducing risks of intraoperative awareness or excessive suppression by tracking proximity to the inflection point.

## 5 Conclusion

By linking baseline neural complexity to a reproducible inflection point in CECP, we provide a mechanistic framework for characterizing divergent responses to anesthesia, bridging empirical EEG dynamics with dynamical systems theory.

## Data Availability

The Chennu dataset is available online at https://www.repository.cam.ac.uk/items/b7817912-50b5-423b-882e-978fb39a49df
The RecCognition dataset used in the present study is available upon reasonable request to the authors

https://www.repository.cam.ac.uk/items/b7817912-50b5-423b-882e-978fb39a49df

## 6 Acknowledgments

CM received funding from Fondation pour la recherche médicale (SPF202409019374). SBM is supported through the Natural Sciences and Engineering Research Council (NSERC) of Canada Discovery Grant (RGPIN-2023-03619); and a Canada Research Chair of Consciousness and Personhood Technology.

