## Supplementary Materials for "Reframing “Paradoxical” Excitation: Disentangling EEG Complexity and Entropy Reveals Resting State Dynamics Associated with Propofol Susceptibility"

### Supplemental Methods

#### Chennu Dataset Participant Classification

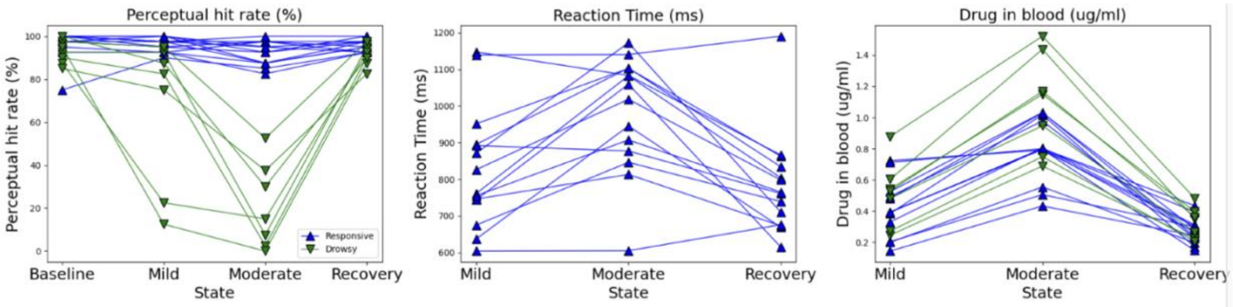


**Supplementary Figure S2. Behavioral Grouping of Chennu Dataset Across Three Behavioral Measures.** This figure shows the replication of grouping of responsive and drowsy participants as based on a perceptual hit rate threshold of 60% during the moderate sedation state which replicates results from Chennu et al. (2016) and Boncompte at al. (2021). Perceptual hit rate, reaction times, and drug concentration in blood are plotted above across three conditions. Green indicates the drowsy group and blue indicates the responsive group.

#### Phase Space Reconstruction

Both statistical complexity and permutation entropy require phase space reconstruction with embedding parameters. To select appropriate embedding parameters, we used a mutual information approach to estimate time delays and the false nearest neighbor's method to determine embedding dimensions through Neurokit. To ensure comparability across participants, the embedding dimension was fixed to the group-averaged value of $m = 6$, while the individual time delay ($\tau$) was estimated separately for each time series.

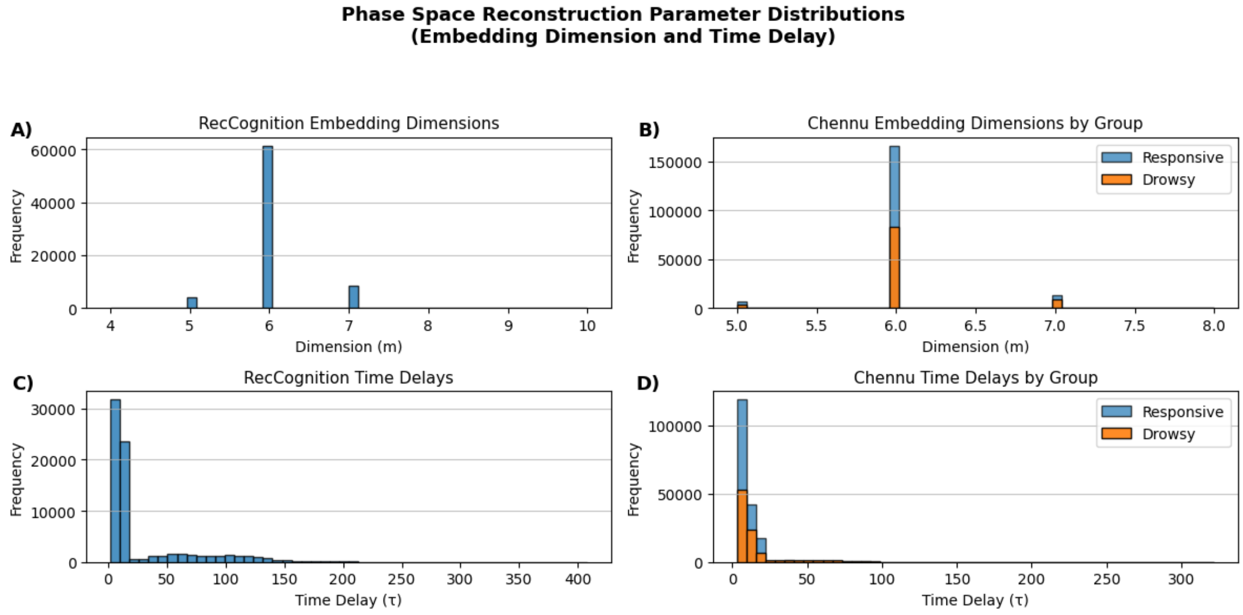


**Supplementary Figure S1. Phase-space reconstruction parameter distributions.** Distributions of embedding dimensions (top row) and time delays (bottom row) computed across all participants, epochs, and channels for both EEG datasets. (A) RecCognition dataset embedding dimensions. (B) Chennu dataset embedding dimensions separated by behavioral group show consistent reconstruction dimensionality across responsive and drowsy participants. The group-averaged dimension, $m = 6$, used for all subsequent analyses. (C) RecCognition time-delay ($\tau$) distribution displays greater variability, reflecting differences in signal autocorrelation structure across participants. (D) Chennu time-delay distributions separated by group.

**RecCognition Dataset Outlier Exclusion**

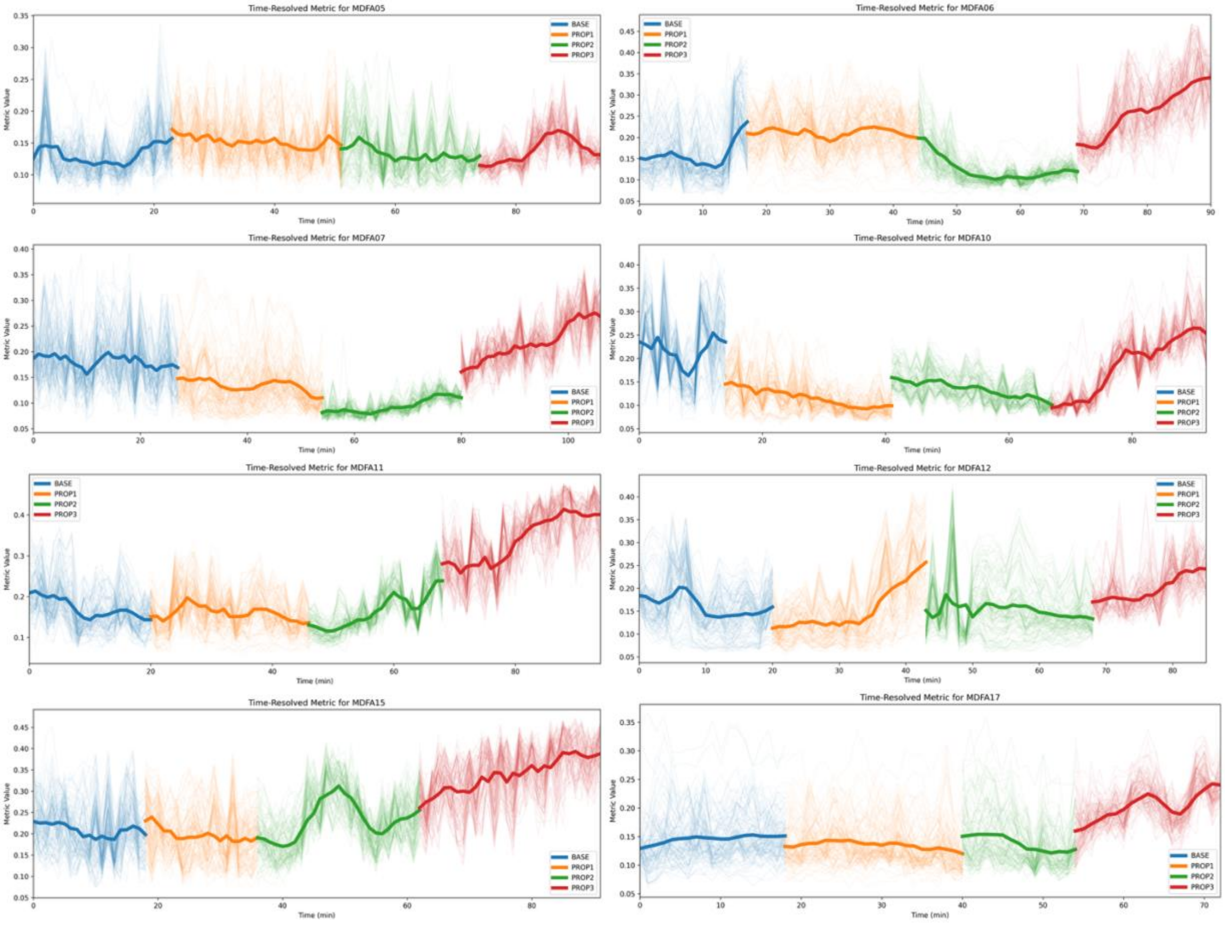


**Supplementary Figure S3. Time resolved complexity for each participant with outlier identification.** For each participant**,** each state's (baseline (blue), propofol 1 (orange), propofol 2 (green) and propofol 3 (red)) complexity value is plotted per epoch (10 second interval) across time. The bold lines represent whole brain average and less opaque lines represent channels. MDFA15 was identified as an outlier due to the increase in propofol 2.

**Simulations**

For the interpretation of empirical EEG signals, we generated various deterministic and stochastic signals to plot on the CECP. We created periodic signals (regular oscillations) by summing sine waves of various frequencies (1–60 Hz) to represent deterministic oscillations. We generated colored noise with varying spectral slopes (from −2 to +2) using filtering of Gaussian white noise. We created Fractional Brownian motion (fBm) signals using the Davies–Harte method from the fbm package[1], simulating time series with Hurst exponents of 0.3, 0.5, and 0.7.

**Complexity-Entropy Causal Plane**

The CECP quantifies the degree of organization in respect to randomness. Importantly, regions of the plane with high complexity and intermediate entropy are associated with causal emergence—structured dynamics that arise from endogenous processes. Yet, it remains an open question whether exogenous perturbations, such as pharmacological sedation, might also induce organized dynamics or shift the brain into low-entropy, high-complexity regimes. This makes the CECP well-suited to assess shifts in the brain's dynamical regime during sedation and anesthesia, where transitions between resting-state consciousness and pharmacologically driven states may reflect changes in complexity profiles.

We projected boundaries of the minimum and maximal complexity along with boundaries of the chaotic region which were estimated by fitting a nonlinear function to empirical maxima on the CECP and used to identify non-linear stochastic regions replicating the CECP form Zanin & Olivares (2020)[2]. Additionally, we computed a representative maximum and minimum complexity using the selected embedding parameters and fixed signal length with Ordpy.

### Supplemental Results

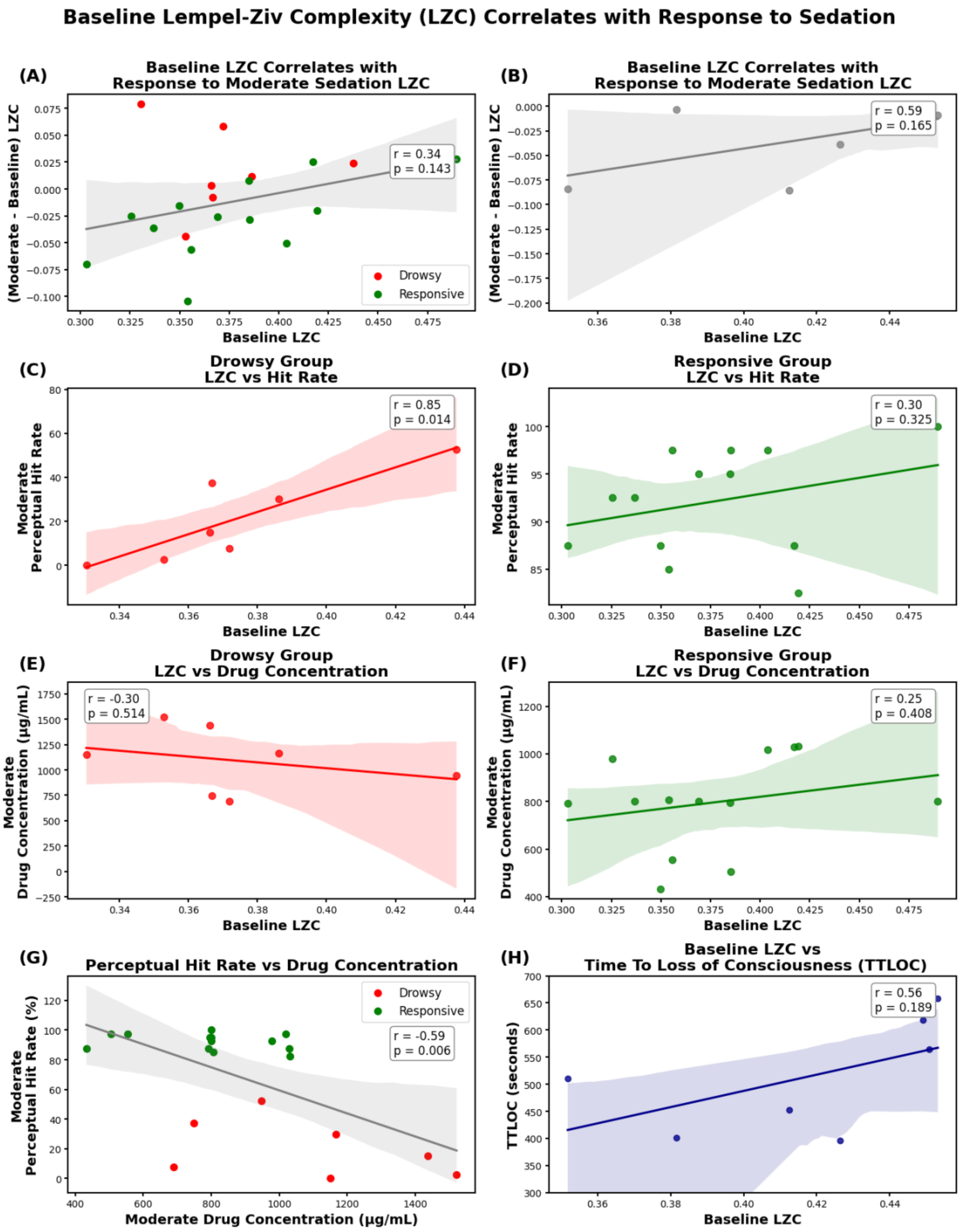

**Supplementary Figure S2. Baseline Lempel–Ziv Complexity is associated with neural and behavioral sensitivity to sedation.** Analogous to Figure 2, this analysis reproduces all correlations using Type I complexity (Lempel–Ziv Complexity) instead of statistical complexity (Type II). (A–B) In both the Chennu and RecCognition datasets, higher baseline LZC showed similar trends with the magnitude of neural response to propofol, though the associations were generally weaker. (C–D) Baseline LZC was modestly related to behavioral responsiveness, with the drowsy group showing a positive trend between baseline complexity and perceptual hit rates under sedation. (E–F) As in the main analysis, baseline LZC was not significantly correlated with drug concentration, indicating that variability in sedation response primarily reflects intrinsic neural state rather than pharmacokinetics. (G) Same as Figure 2G. (H) Shaded regions indicate 95% confidence intervals of linear fits; reported statistics are Pearson correlation coefficients.

##### 3.3.1. Mapping the response to anesthesia in a complexity-entropy causal plane reveals a critical inflection point: Situating simulated signals.

The placement of simulated signals within the CECP establishes a continuum from highly structured, low-entropy periodic oscillations to high-entropy, low-complexity white noise, providing a comparative reference for interpreting the EEG trajectories observed during anesthesia induction.

Regular oscillations, generated from sinusoidal signals at varying frequencies, occupy the low-entropy, low-complexity region, characteristic of deterministic systems with minimal randomness. In contrast, colored noise processes, modulated by spectral slope variations, distribute along a high complexity and high entropy, reflecting nonlinear stochasticity with varying degrees of temporal correlations. Similarly, Fractional Brownian motion, parameterized by different Hurst exponents, exhibits increasing entropy and decreasing complexity, indicative of memory-dependent random processes. At the extreme, white noise, defined by maximal entropy and minimal complexity, serves as a benchmark for a true random process.

### Supplemental Discussion

##### Both Type I and Type II complexity capture paradoxical excitation during mild sedation

Whereas paradoxical effects were present at mild sedation across datasets and types of complexity, the paradoxical effect vanishes with higher levels of propofol. During deep sedation, measures rebound—LZC decreased while SC increased—indicating a reconfiguration of neural dynamics during loss of consciousness. Although SC was not significantly different between groups at moderate sedation, the Mann-Whitney U statistic indicated a larger rank separation between groups than for LZC, suggesting SC may capture paradoxical excitation more robustly in larger cohorts.

Type II complexity represents a broader class of measures capturing the emergence of structured patterns[3], with prior work showing Type I and II can diverge under anesthesia[4]. Li et al., (2022)[5] review complexity measures across temporal, spatial, and spatiotemporal domains under anesthesia, such as microstate dynamics[6].

##### Baseline complexity determines susceptibility to propofol

This suggests that susceptibility to propofol and the presence of paradoxical excitation are predetermined by pre-anesthetic neural dynamics. In other words, sedation drives all brains toward a noisy, unstable regime, but the initial dynamic regime before drug exposure (i.e., the baseline complexity) dictates the path through it. Individuals who start closer to the instable inflection point (i.e. low-SC)  are rapidly suppressed and show minimal or absent paradoxical reactions. Meanwhile, individuals whose initial brain dynamics are further away from the inflection point (i.e. high-SC) cross the instable inflection point before showing suppression and thus show paradoxical excitation before eventual collapse.

The concept of neural inertia, the brain's tendency to resist transitions in arousal state, is highly relevant here. Luppi et al. (2021) describe it as hysteresis between anesthetic induction and emergence, where the prior brain state influences responsiveness[7]. Our results suggest that baseline complexity reflects a state–dependent inertia, indicating whether the brain transitions into paradoxical excitation or straight into suppression under sedation.

**Mapping neural trajectories on the CECP uncovered an instable inflection point**

Mechanistically propofol may act in two phases. At low–moderate doses, propofol potentiates $GABA_A$ currents within inhibitory networks, driving interneuron antisynchrony and beta-band paradoxical excitation that destabilizes ongoing temporal structure. At higher doses, strengthened inhibition imposes deterministic constraints entraining thalamocortical circuits into coherent frontal alpha rhythm and subsequently burst-suppression[8-12]. The inflection point we observed appears to mark the transition between these two regimes, the collapse of endogenous temporal structure and the onset of exogenously imposed causal constraints. In both datasets, trajectories converged on an inflection point in the CECP (Figure 4), where minimal complexity rebounds, and trajectories bifurcate between excitation and suppression.

**Future directions**

Beyond anesthesia, the framework may also be valuable for the evaluation of patients in a disorder of consciousness. In previous research from our group, we described a case series of unresponsive patients who showed a paradoxical response to propofol[13]. Likewise, mounting evidence describes paradoxical effects of the GABA-ergic drug Zolpidem to unresponsive patients[39-42]. Most importantly, response to Zolpidem is extremely heterogenous with paradoxical therapeutic effect being observed in only 5-12 % of patients[14,15]. The potential use of the CECP framework to investigate endotypes of drug responders should be topic of future research.
